# Smart Pooling: AI-powered COVID-19 testing

**DOI:** 10.1101/2020.07.13.20152983

**Authors:** María Escobar, Guillaume Jeanneret, Laura Bravo-Sánchez, Angela Castillo, Catalina Gómez, Diego Valderrama, Maria F. Roa, Julián Martínez, Jorge Madrid-Wolff, Martha Cepeda, Marcela Guevara-Suarez, Olga L. Sarmiento, Andrés L. Medaglia, Manu Forero-Shelton, Mauricio Velasco, Juan Manuel Pedraza-Leal, Silvia Restrepo, Pablo Arbelaez

## Abstract

**Background:** COVID-19 is an acute respiratory illness caused by the novel coronavirus SARS-CoV-2. The disease has rapidly spread to most countries and territories and has caused 14·2 million confirmed infections and 602,037 deaths as of July 19^th^ 2020. Massive molecular testing for COVID-19 has been pointed as fundamental to moderate the spread of the disease. Pooling methods can enhance testing efficiency, but they are viable only at very low incidences of the disease. We propose Smart Pooling, a machine learning method that uses clinical and sociodemographic data from patients to increase the efficiency of pooled molecular testing for COVID-19 by arranging samples into all-negative pools.

**Methods:** We developed machine learning methods that estimate the probability that a sample will test positive for SARS-Cov-2 based on complementary information from the sample. We use these predictions to exclude samples predicted as positive from pools. We trained our machine learning methods on samples from more than 8,000 patients tested for SARS-Cov-2 from April to July in Bogotá, Colombia.

**Findings:** Our method, Smart Pooling, shows efficiency of 306% at a disease prevalence of 5% and efficiency of 107% at disease a prevalence of up to 50%, a regime in which two-stage pooling offers marginal efficiency gains compared to individual testing (see Figure 1). Additionally, we calculate the possible efficiency gains of one- and two-dimensional two-stage pooling strategies, and present the optimal strategies for disease prevalences up to 25%. We discuss practical limitations to conduct pooling in the laboratory.

**Interpretation:** Pooled testing has been a theoretically alluring option to increase the coverage of diagnostics since its proposition by Dorfmann during World War II. Although there are examples of successfully using pooled testing to reduce the cost of diagnostics, its applicability has remained limited because efficiency drops rapidly as prevalence increases. Not only does our method provide a cost-effective solution to increase the coverage of testing amid the COVID-19 pandemic, but it also demonstrates that artificial intelligence can be used complementary with well-established techniques in the medical praxis.

**Funding:** Faculty of Engineering, Universidad de los Andes, Colombia.

**Research in context:** *Evidence before this study:* The acute respiratory illness COVID-19 is caused by severe acute respiratory syndrome coronavirus 2 (SARS-CoV-2). The World Health Organization (WHO) labeled COVID-19 as a pandemic in March 2020. Reports from February 2020 indicated the possibility of asymptomatic transmission of the virus, which has called for molecular testing to identify carriers of the disease and prevent them from spreading it. The dramatic rise in the global need for molecular testing has made reagents scarce. Pooling strategies for massive diagnostics were initially proposed to diagnose syphilis during World War II, but have not yet seen widespread use mainly because their efficiency falls even at modest disease prevalence. We searched PubMed, BioRxiv, and MedRxiv for articles published in English from inception to July 15^th^ 2020 for keywords “pooling”, “testing” AND “COVID-19”, AND “machine learning” OR “artificial intelligence”. Early studies for pooled molecular testing of SARS-CoV-2 revealed the possibility of detecting single positive samples in dilutions of samples from up to 32 individuals. The first reports of pooled testing came in March from Germany and the USA. These works suggested that it was feasible to conduct pooled testing as long as the prevalence of the disease was low. Numerous theoretical works have focused only on finding or adapting the ideal pooling strategy to the prevalence of the disease. Nonetheless, many do not consider other practical limitations of putting these strategies into practice. Reports from May 2020 indicated that it was feasible to predict an individual’s status with machine learning methods based on reported symptoms.

*Added value of this study:* We show how artificial intelligence methods can be used to enhance, but not replace, existing well-proven methods, such as diagnostics by qPCR. We show that in this fashion, pooled testing can yield efficiency gains even as prevalence increases. Our method does not compromise the sensitivity or specificity of the diagnostics, as these are still given by the molecular test. The artificial intelligence models are simple, and we make them free to use. Remarkably, artificial intelligence methods can continuously learn from every set of samples and thus increase their performance over time.

*Implications of all the available evidence:* Using artificial intelligence to enhance rather than replace molecular testing can make pooling testing feasible, even as disease incidence rises. This approach could make pooled testing an effective tool to tackle the disease’s progression, particularly in territories with limited resources.

## 2 Introduction

COVID-19 is an acute respiratory illness caused by the novel coronavirus SARS-CoV-2 [1, 2]. The disease has rapidly spread to most countries and territories and has caused 14·2 million confirmed infections and 602,037 deaths as of July 19^th^ 2020 [3]. Clark *et al*. [4] estimated that 1·7 billion people, which corresponds to 22% of the global population, are at risk of developing severe COVID-19. This susceptible fraction of the population have at least an underlying condition related to the increased risk of the disease.

**Fig 1.**
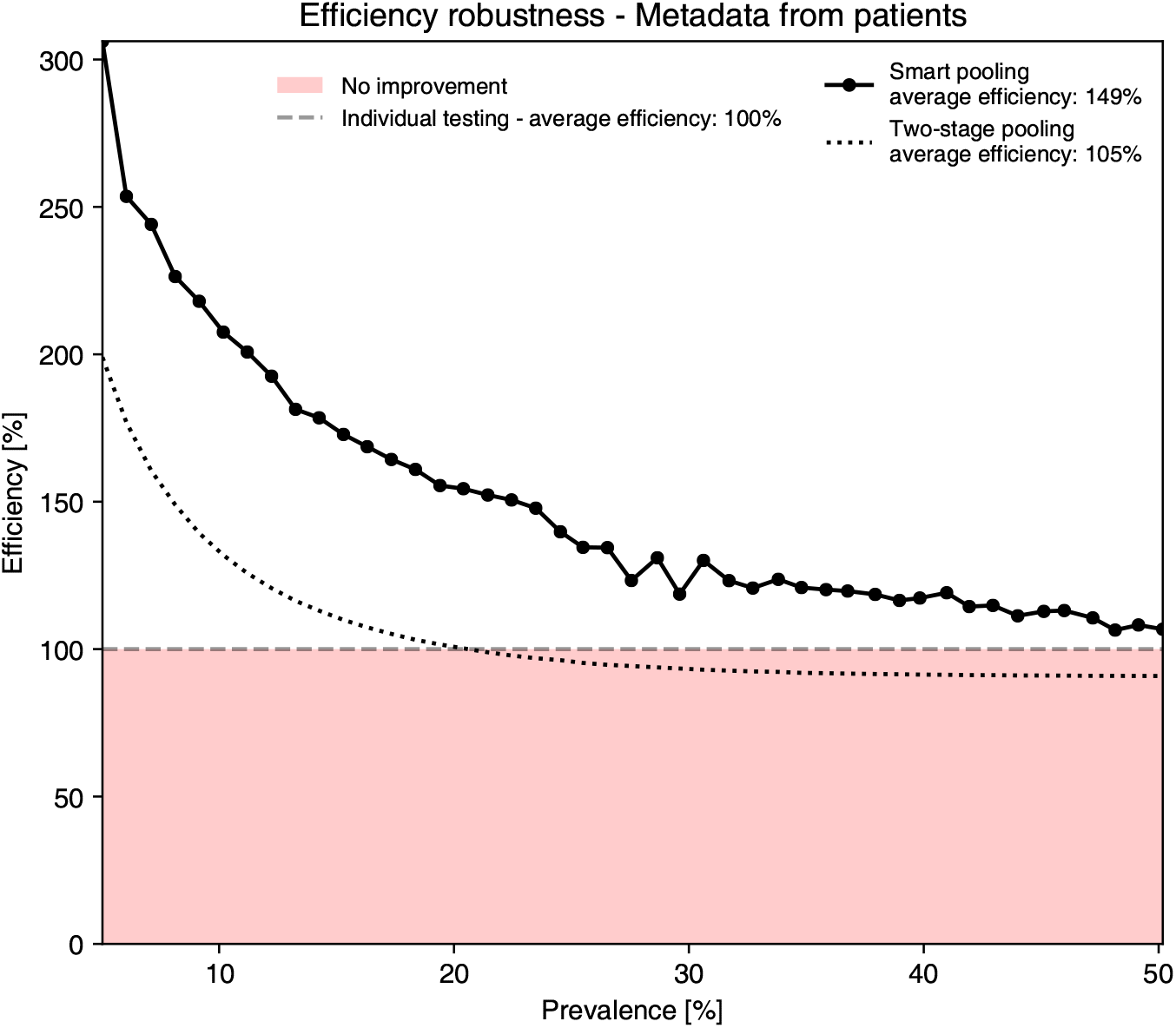
Efficiency of Smart Pooling compared to standard testing methods on Patient Dataset. Smart pooling achieves higher efficiencies than two-stage pooling and individual testing for prevalences of the disease of up to 50% on the Patient Dataset. The average efficiency measures the overall efficiency of each method in the complete prevalence range.

Countries are rushing to implement massive testing to identify and isolate carriers to tackle the spread of the disease. Nevertheless, massive testing is costly and has increased the demand for scarce reagents. However, there is still a global need to make testing more accessible to larger populations. Thus, strategies to enhance the efficiency of testing, that is, the number of people that can be tested with the same amount of test kits, are urgent.

Pooling methods, which were initially proposed by Dorfman to diagnose syphilis among the US military during World War II [5], allow to test several patients with fewer reagents by combining their samples in a single test tube. Different pooling strategies, such as two-stage pooling [5] and matrix pooling [6], offer more significant efficiency gains for different combinations of test sensitivity and disease prevalence. Pooling has not only been used for diagnosis but also in genetic sequencing [7].

Empirical tests have confirmed the ability of pooled analysis to reliably detect SARS-CoV-2 in a pool comprising one positive specimen and up to 31 negative specimens [8, 9]. Additionally, preliminary trials show that pools of 10 specimens can increase efficiency without strongly compromising sensitivity [10, 11]. Non-adaptive pooling scheme demonstrates an increase in the efficiency for low prevalences in in-vitro experiments [12]. Given sufficient sensitivity for the test, pooling methods are effective when the prevalence of the disease is low so that the probability of all samples in a pool being negative is high, but fails as the prevalence increases, as shown in Figure 2a.

**Fig 2.**
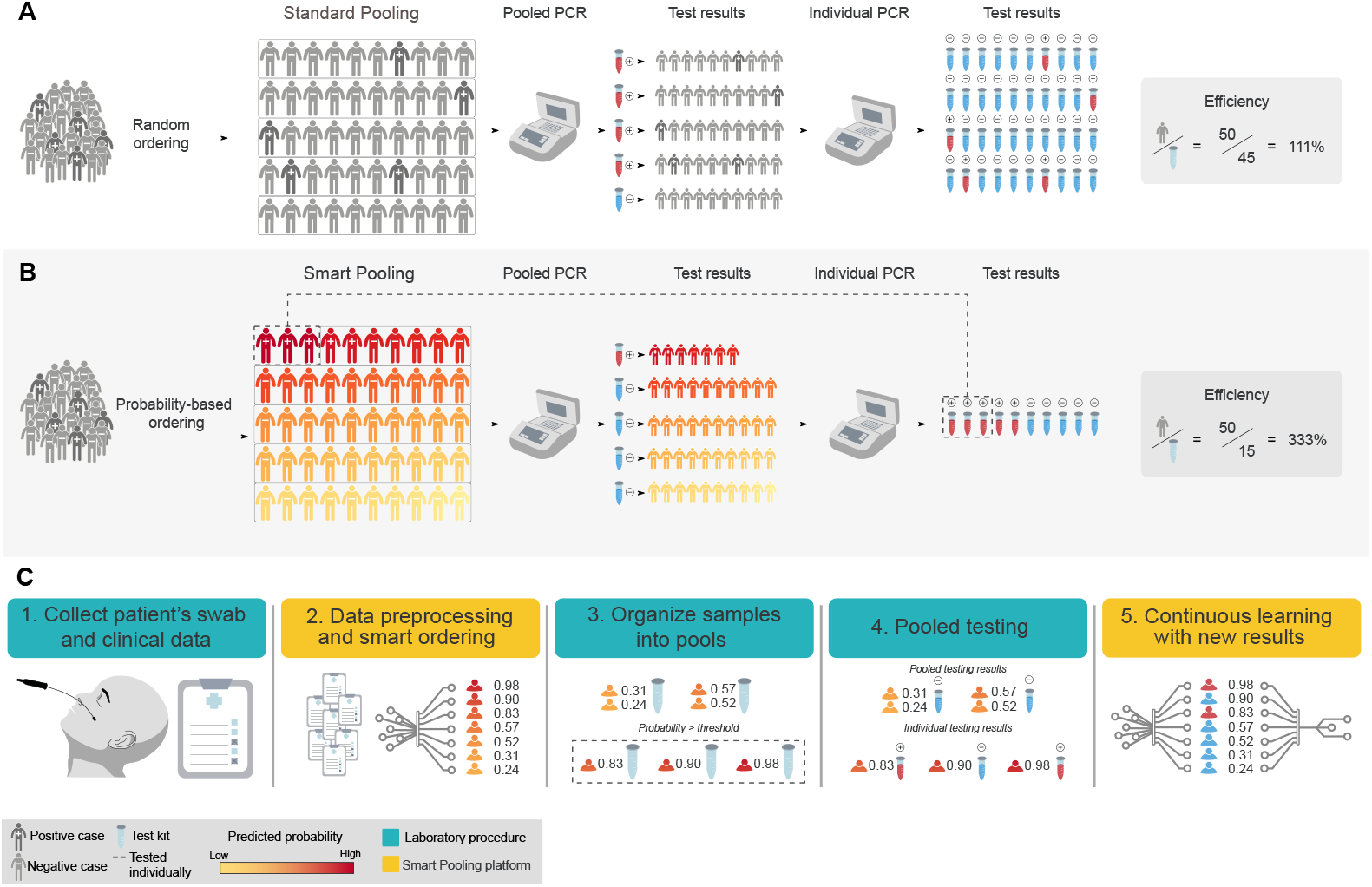
Smart Pooling makes pooling methods efficient even at high disease prevalences. **A** In standard two-stage pooling methods, samples are pooled randomly. When the outcome of a pooled test is negative, all the samples in it are labeled as negative. When the outcome is positive, all samples are tested individually. As prevalence increases, the efficiency of pooling without a priori information drops rapidly and makes the strategy unviable, mainly because the probability of having at least one positive sample in a pool increases. **B** Smart Pooling tackles this problem by ordering samples according to the predicted probability of yielding a positive result in the test. If the probability of a sample surpasses the defined threshold, this sample goes directly to individual testing. On the contrary, if a sample has a lower probability, it goes to pooled testing. We arrange samples into pools with similar probability, maximizing the probability of being all-negative. **C** *The Smart Pooling pipeline*. Samples and data are collected from patients. The Smart Pooling model processes these data and returns an arrangement with the probability that each sample tests positive. We compare this probability to a threshold and, if it is greater than the threshold, we select the sample for individual testing. In the laboratory, samples are tested individually or pooled based on the defined arrangement. Subsequently, samples from positive pools are tested individually. Finally, the diagnostic outcome of each sample is fed to the Smart Pooling platform. This process enlarges the dataset and allows for continuous learning.

Here we present Smart Pooling, a machine learning method that enhances the efficiency of pooling testing strategies. Smart Pooling exploits clinic and sociodemographic information of samples to estimate their probability of testing positive for COVID-19. As Figure 2b shows, our method uses these probabilities to arrange all samples into pools that maximize the probability of yielding a negative result within the most number of pools. That is, we group samples such that positive samples are excluded from the pooling process and are evaluated in individual tests, thus reducing the number of tests used for the same amount of samples.

Machine learning and regression models have been used to classify the novel pathogen from its genetic sequencing [13], to support diagnosis from CT scans [14], to assist clinical prognosis of patients [15], and to forecast the evolution of the pandemic [16]. Regression models trained with patient-reported symptoms and laboratory test results have been used to predict infection from symptomatology [17]. These strategies risk compromising sensitivity and confidence. In contrast, Smart Pooling does not seek to replace current molecular testing but assist it by improving its efficiency.

Complementary information from samples, such as microscopic inspections, has been effectively used to guide pooling methods in detecting malaria [18]. In the epidemiological context of the COVID-19 pandemic, testing laboratories may have clinical and sociodemographic data for each individual. These data could be exploited to estimate the probability of a patient yielding a positive result [19]. Thus, an informed guess could be made to exclude a sample from a pool. Furthermore, new studies [20] show that to reduce the transmission of SARS-CoV-2, it is desired to combine isolation and tracing strategies. The number of tests enhances these tracing strategies as the exploration of wider contact networks is possible.

Smart Pooling is easily adaptable to any pooling procedure. To use Smart Pooling as a tool for the desired pooling strategy, we propose a five-step pipeline between the laboratory procedures and the Smart Pooling analysis. Figure 2c illustrates this process. First, the laboratory acquires samples and sociodemographic metadata from patients. Secondly, the Smart Pooling platform processes metadata to provide an ordering of the patients into pools or send them to individual testing when there is a high probability of yielding a positive result. Thirdly, samples are pooled according to this ordering in the laboratory. Then, molecular tests are run individually or on the ordered pools until there is a diagnosis for each sample. Lastly, the laboratory feeds the results of the tested samples into the Smart Pooling platform to continuously improve the model.

## 3 Methods

### 3.1 Dataset

The data used in our work corresponds to the molecular tests conducted by Universidad de Los Andes for the Health Authority of the city of Bogotá, Colombia. To construct the dataset, we tested samples individually following the Berlin Protocol [21] (for dates before April 18^th^) and the protocol for the U-TOP™ COVID-19 Detection kit from Seasun Biomaterials [22]. Our dataset consists of two different groups, representing the diversity of available data.

#### 3.1.1 Patient Dataset

We organized the first group of data according to the outcome of each molecular test. We also had access to the patient’s: sex; age; date of onset of symptoms; date of the medical consultation; initial patient classification; information about the patients’ occupation, specifying if they are health workers; patients’ affiliation to the health system; data about patients’ travels, pointing if they recently had international or domestic travels; comorbidities; symptoms; and if they had had contact with a confirmed or suspected COVID-19 case. Samples were collected from April 18^th^ to July 15^th^ 2020, comprising 2,068 samples. The patients’ median age was 36 years, and the age range was between 0 and 93 years. 1,142 patients were male (55%) and 926 (45%) were female. The additional information followed the protocol developed by the Colombian Public Health Surveillance System - Sistema Nacional de Vigilancia en Salud Publica (SIVIGILA).

#### 3.1.2 Test Center Dataset

In the second group of data, we organized data according to the test center that collected the sample. We had access to metadata from the test centers, such as their location and name. Samples were collected from April 6^th^ to May 25^th^ 2020, with a total of 7,162 samples from 101 test centers.

### 3.2 Training of the predictors

#### 3.2.1 Dataset division

To experimentally validate the performance of our proposed models and descriptors, we followed a two-fold cross-validation scheme for the *Patient Dataset*, where we used one data fold for training and the held-out fold for evaluating the model. We obtained the final result by evaluating each fold on the corresponding model and then aggregating the predictions to calculate the performance metrics in the entire dataset. We created the folds using a stratified strategy, where each fold had the same percentage of positive and negative samples, as well as a similar distribution of other factors, such as age, sex, or symptoms. For the *Test Center Dataset*, we divided the data into a training split containing all the samples until May 7^th^, and a held-out test split for evaluating the performance of the model with the remaining samples. We further divided the training data into two disjoint subsets: training and validation. We selected the validation subset such that we predicted as many dates as those in the test set. The maximum number of dates to predict after May 7^th^ was five.

#### 3.2.2 Evaluation

##### Quantitative evaluation

We measured the efficiency of our models in silico in terms of the number of tests used when employing the model’s output as the criterion for Smart Pooling. We calculated the efficiency as the fraction between the number of samples and the total amount of tests used, following a two-stage pooling strategy. First, we sorted the individual tests based on the predicted prevalence by decreasing order. Then, we grouped the samples following the original order and the pool size. Finally, the total number of tests corresponds to the number of pools in the first step added to the individual samples from the second step, that is, samples from the pools that were positive in the first step. In our experiments we report the efficiency as a percentage.

#### 3.2.3 Model training

We trained a machine learning method independently for each dataset. We used the AutoML from H2O library [23] to explore multiple machine learning models in the validation sets. Below we explain the details for training at each dataset.

##### Patient Dataset

In this dataset, we trained our methods to classify the test of a given patient into a positive or negative sample for COVID-19. Then, we used the probability of the sample being positive, as predicted by the model, to organize the tests into pools and calculate the efficiency. For these experiments, we defined a descriptor in which each feature dimension corresponds to a variable from the patient and location information, previously defined in section 3.1.1. The best model for this task was a Gradient Boosting Machine (GBM) [24] with 30 trees, a mean depth of 9, a minimum of 12 leaves, and a maximum of 22 leaves.

Initially, we split the data into two sets to perform cross-validation. To establish similar conditions between each subset of the dataset, we randomly chose the total number of positive and negative samples such that the number of data points across each subset remained similar. Secondly, we trained our model over one subset, say the first split, and validate over the one leftover, say the second one. We stored the predicted probabilities for the samples in the second split to use them as the scores. Then, we performed the same process, but we trained with the second split, validated over the first one, and stored the first split’s sample predictions. After completing this process, we organized each sample using the stored probability scores to perform Smart Pooling.

##### Test Center Dataset

The level of granularity of this dataset allowed us to have information on the number of positive and total tests per test center on a given date. However, we do not have daily reports from each test center. Thus, we explicitly modeled the data as a sparse time series. We trained the machine learning methods to predict the fraction of positive tests for a center on the current date. Afterward, we assigned this value as the incidence of each sample from a test center on a given date. We sorted the tests by decreasing incidence, simulated a two-stage pooling protocol, computed the number of used tests, and calculated the efficiency of Smart Pooling. The best model for this task was a Gradient Boosting Machine (GBM) [24] with 50 trees, a constant depth of 5, a minimum of 12 leaves, and a maximum of 29 leaves.

To predict the incidence for a date in the validation or test set, we defined a descriptor calculated from the available training data for each testing center. In the features, we included the cumulative tests of each institution up to every date within the time series *i.e*. the institution prevalence, and the total number of tests from all the institutions at the corresponding date. To include the temporal information, we defined as features the date in YY-MM-DD format and the relative date in the number of days since the first date on the time series. Additionally, we created a variable that we named gap, which encodes the distance between two consecutive entries from the same test center. In particular, during the testing stage, the gap encodes the distance between the last training date and the date to predict. The gap provided the model with an estimate of the analyzed time window.

As such, we compute the descriptor’s features by analyzing the relative differences between variables on the last known date and those from the training days in the current gap. These gap features comprise the cumulative number of tests, total tests, number of positive samples for each institution at the last available date, and the corresponding incidences. We complemented the descriptor with features that indicate the number of days, relative to the current prediction date, from the first *n* number of positive tests in each test center. For our experiments we defined *n* = 1, 5, 10, 100, 500, 1000. The rationale behind this feature was to provide the model with a temporal encoding of the incidence’s evolution. We adapted these features from a public kernel from Kaggle’s COVID-19 Global Forecasting Challenge ^1^.

We performed the training of our models in two stages. In the first training phase, we used the validation subset’s performance as the criterion to select the best model. We removed the test centers that did not have sufficient dates for the validation split but included them back for the second training phase. Lastly, we obtained our final model by retraining the best model using all available training data and evaluated it on the test split.

The key idea of Smart Pooling is maintaining *p* artificially low, even under scenarios with a high overall prevalence of COVID-19, by reordering the samples according to a priori estimates of prevalence, before the pooling takes place. We demonstrate efficiency gains at simulated prevalences of up to 25% and 50% for the *Test Center Dataset* and *Patient Dataset*, respectively, which are considerably higher than those applicable for even optimal standard pooling methods.

## 4 Results

### 4.1 Smart Pooling increases testing efficiency by excluding positive samples from pools

We identify that using complementary information to arrange pools can improve the efficiency of testing. We do this by training a machine learning algorithm to predict the probability that a sample will test positive for COVID-19 based on clinical and sociodemographic data. Testing efficiency increases by using individual testing on high probability samples and arranging the remaining samples into pools simulating a two-stage pooling protocol. Figures 1 and 3 show that for disease prevalences ranging from 0% up to 50% and from 0% up to 25%, respectively, Smart Pooling’s efficiency outperforms the simulated efficiency obtained with Dorfsman’s two-stage pooling and individual testing.

**Fig 3.**
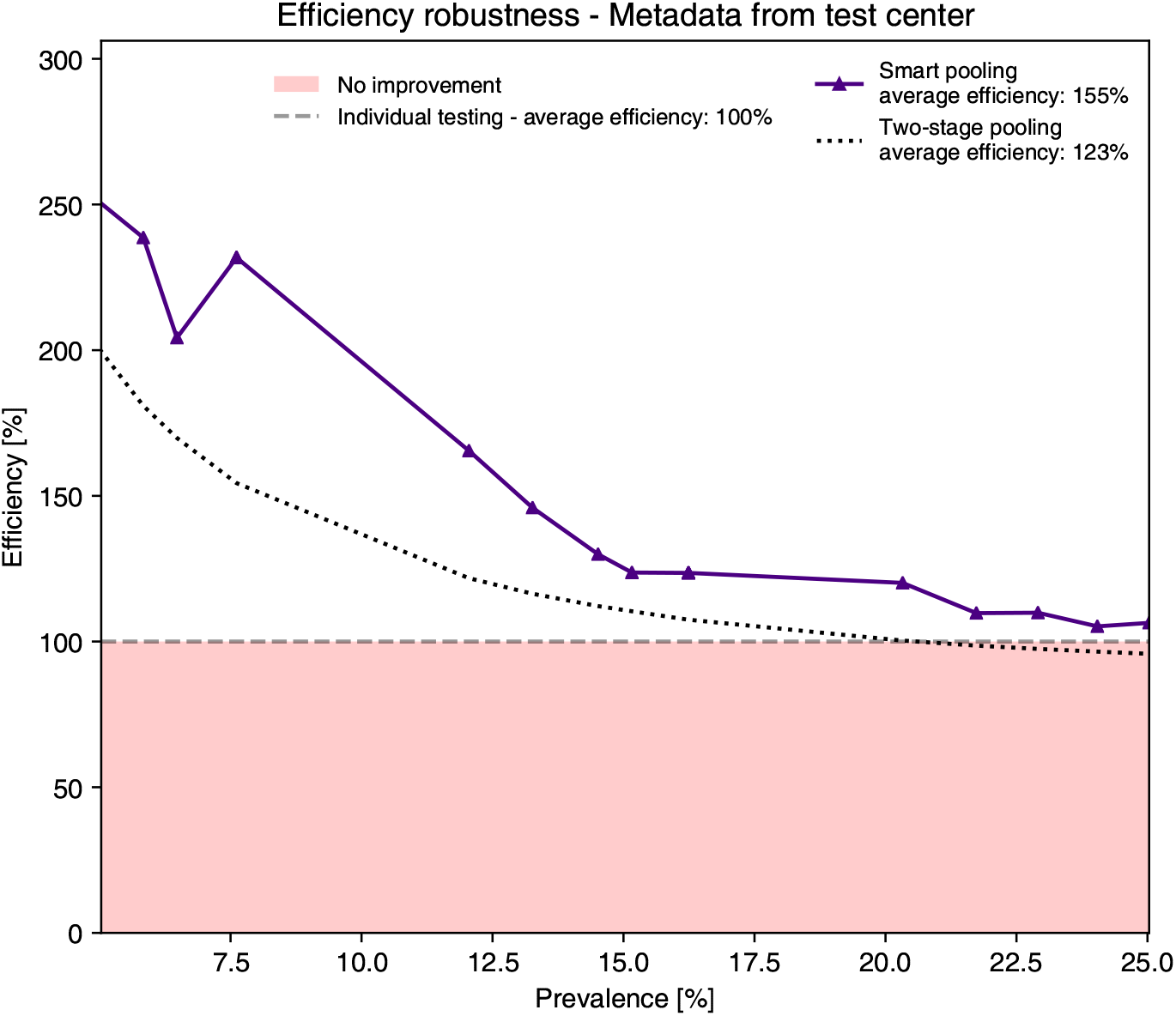
Efficiency of Smart Pooling trained with the coarse metadata from the Test Center Dataset. Despite not having detailed patient metadata, Smart pooling is still capable of producing higher efficiencies than two-stage pooling for prevalence of the disease of up to 25%. The average efficiency measures the overall efficiency of each method in the complete prevalence range.

Our computational experiments show that, regardless of the level of granularity of the data available for performing Smart Pooling (Figures 1 and 3), efficiency gains are obtained for all simulated prevalences up to 25% for the *Test Center Dataset* and for the *Patient Dataset* up to 50%. For instance, with Smart Pooling at a prevalence of 10% and an efficiency around 200%, even with coarse data in the test center approach, the estimated number of patients that could be tested with the same number of tests is doubled compared to individual testing. This shows that Smart Pooling could be viable in various settings and does not depend on the availability of rich complementary data.

Figure 4 depicts the visualized predictions from our machine learning models trained with the patients’ metadata. We rank samples according to the confidence of the model for predicting a sample as positive. Compared to the random ordered samples, for Smart Pooling’s predictions based on the patients’ dataset, most of the positive samples on top of the arrangement. This figure illustrates the working principle of Smart Pooling: it enhances efficiency by artificially reducing the incidence in the samples sent to pools, by sending the samples most likely to test positive to individual testing.

**Fig 4.**
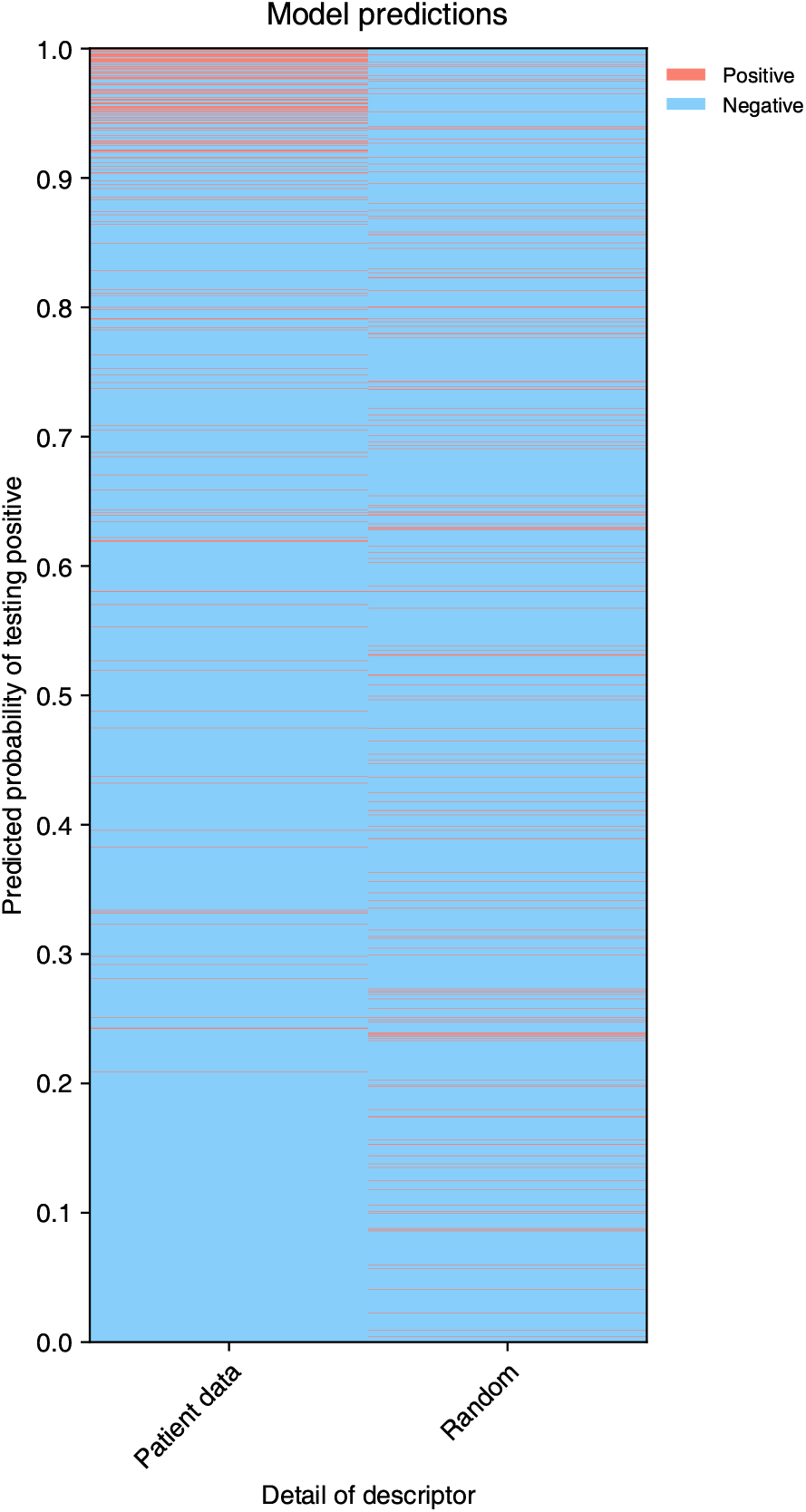
Visualization of Smart Pooling model predictions trained with patient level metadata. Smart Pooling arranges samples by the probability of testing positive based on the predictions made by each model, and we compare this proposed order to a random arrangement.

The machine learning model takes advantage of complementary data to compute the probability that a sample results positive. For the *Patient Dataset*, this could come from reported symptoms or the knowledge that the patient belongs to a particular group (for instance, being a health worker). For the *Test Center Dataset*, the machine learning model could be exploiting underlying correlations in the samples [25, 26]. In other words, Smart Pooling could be seen as the assembly of pools with correlated samples where the independence assumption in the underlying binomial distribution of Dorfman’s protocol does not longer hold (within the pool of *correlated* samples) [26]. Samples in this dataset were acquired during strict measures limiting mobility in the city of Bogotá. It is likely that people tested at the same center shared epidemiological factors, like visiting the same markets, sharing public transport, or being in the same hospital. The model could also be learning the different probability distributions of samples being positive in different parts of the city.

Smart Pooling’s use is not limited to two-stage pooling; it can be coupled with multiple pooling strategies and improves efficiency regardless of the strategy. Figure S1 shows the effect of using Smart Pooling with a fixed pool size of 10 and an adaptive pooling strategy based on the optimal strategies explored in the following section. Both of these alternatives are more efficient than two-stage pooling and individual testing when coupled with Smart Pooling. Our simulations show that using adaptive pooling strategies offers efficiency gains than fixed-size pooling for higher prevalences (*p >* 15%). Ultimately, the pooling strategy used in practice should be determined by the resources available at the testing laboratory.

### 4.2 Pooling strategies

#### 4.2.1 Two-stage pooling protocols

By a two-stage pooling protocol, we mean a procedure that, given a set of samples, is combined into pools using specific criteria. Once the pools are defined, we carry out two stages. In the first stage, we test each pool for disease using qPCR. There are two possible outcomes: (i) The pooled sample is *negative*, which implies that *all* individual samples within the pool are negative; or (ii) The pooled sample is *positive*, which implies that *at least one* individual sample within the pool is positive. If the test result of the first stage is positive, then it is necessary to proceed to the second stage. In the second stage, each sample is individually tested, thereby finding the infected samples. More importantly, note that if the result of the first stage is negative, by performing a single pooled test, we save all the PCR kits required to test the samples individually (except the one used for the pooled test). Crucially, as we will show later, this gain is obtained without any loss in either test sensitivity or specificity.

We explored the following two kinds of pooling protocols:

1. Dorfman’s pooling protocols [5]: Given *m* samples, we make a single pool with all of them. We denote this protocol by *S*_*m*_
2. Matrix pooling protocols [6]: Given a collection of *m* × *n* samples, we place them into a rectangular *m* × *n* array. We create pools by combining samples along the rows and along the columns of this array (for a total of *m* + *n* pools). In the second phase, we test each sample at the intersection of the positive columns and rows individually. We denote this protocol by *S*_*m*×*n*_.

#### 4.2.2 Quantifying pooling efficiency

Pooling efficiency will be our main tool to quantitatively compare different pooling protocols, as it is defined as the number of patients tested per detection kit consumed. Efficiency depends on the prevalence *p* of the sample (defined as the probability that an individual in the population is ill). Assuming independence among patients, pooling efficiency can be easily computed analytically [5, 6]:

1. For Dorfman’s pooling *S*_*m*_ the efficiency is given by

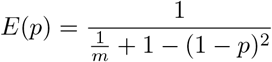
2. For the matrix pooling protocol *S*_*m*×*n*_ the efficiency is given by

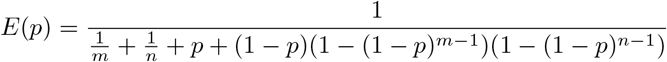

The efficiency functions above show the key property behind pooling in general, and Smart Pooling in particular. At low prevalences, the efficiency can be considerably greater than one. As the prevalence increases, *E*(*p*) decreases, and the efficiency gains become negligible after prevalences around 30%. Figure 5 shows graphically these efficiency gains (and how they vanish) for two-stage pooling.

**Fig 5.**
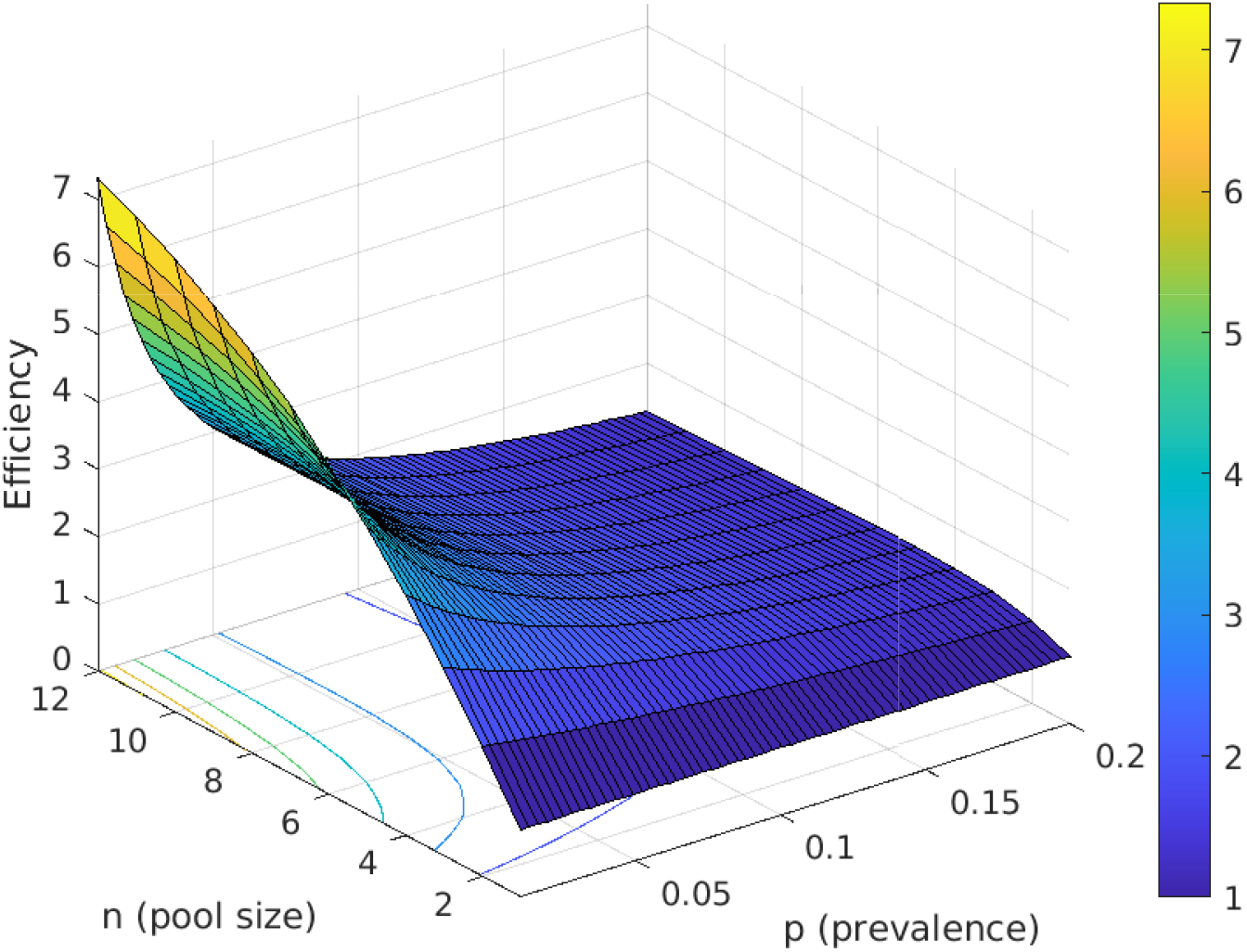
Efficiency of two-stage Dorfman’s pooling as a function of prevalence *p* and pool size *n*

#### 4.3.2 Optimal pooling strategies

What are the best pooling protocols that maximize efficiency for a given prevalence *p*, assuming a fixed bound *c* on pool size? The experimental setup unavoidably constrains the maximum pool size. In the context of COVID-19 we will focus on the cases *c* = 5 and *c* = 10 since these are the most useful in practice (see Section S1.1 for details). Finding the optimal pooling strategies for a given prevalence equals finding the pooling protocol of maximum efficiency by comparing the values of *E*(*p*) for the different protocols, as Mutesa *et al*. also proceed [27]. Figure 6 shows the efficiency curves of the best pooling protocols of the form *S*_*j*_ and *S*_*m*×*n*_ with maximum pool size *c* ≤ 10 and *c* ≤ 5. The optimal two-stage protocols exist (i.e., their efficiencies are the highest among all two-stage pooling protocols, not only among those of the form *S*_*m*_ and *S*_*m*×*n*_). More precisely, table 1 shows the optimal protocols and their respective intervals of optimality when *c* = 10 and *c* = 5 respectively. Even while using optimal protocols, it is clear that the efficiency gains quickly disappear as the prevalence increases (effectively vanishing when *p ≥* 30·66%).

**Table 1.**
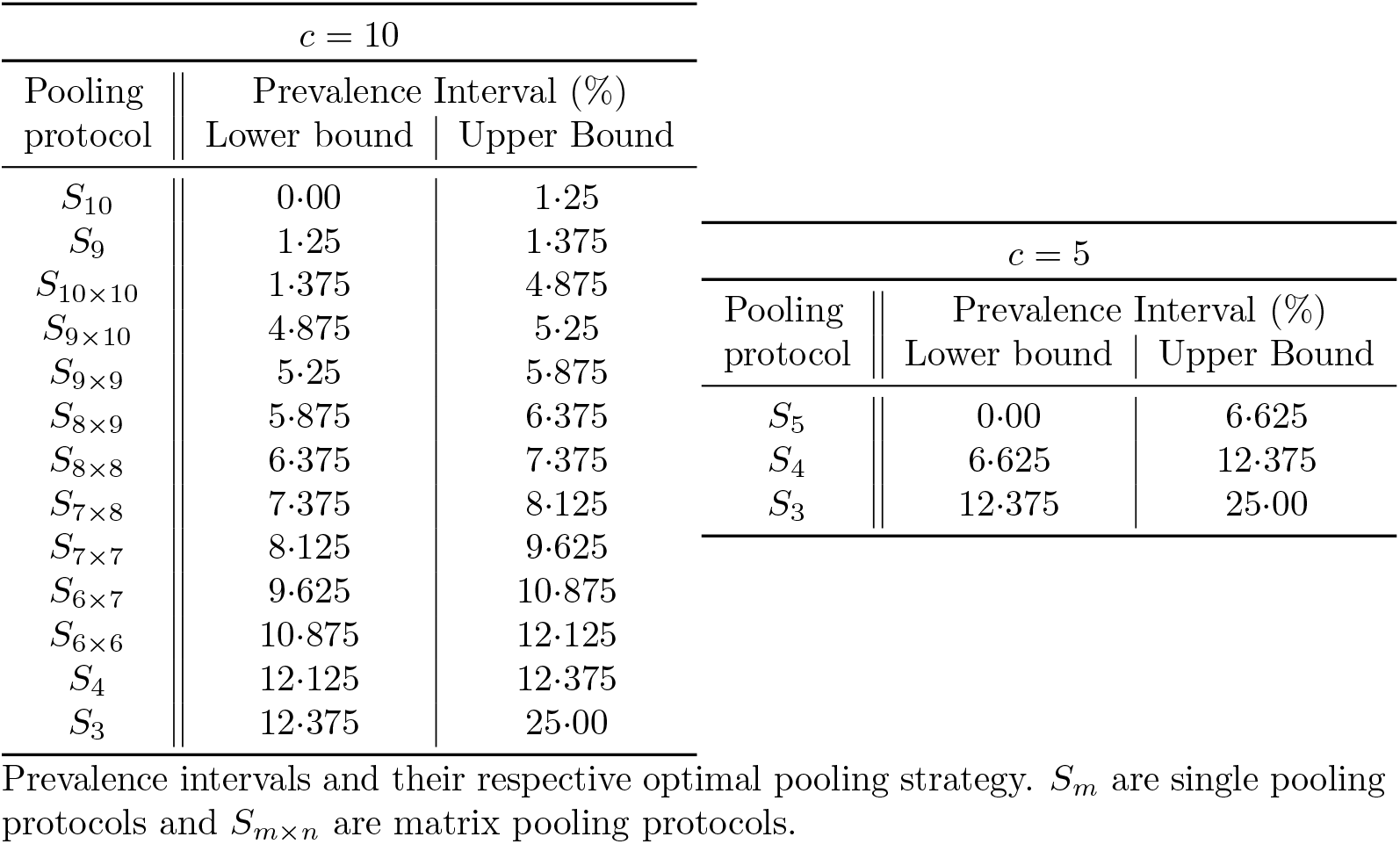
Optimal pooling strategies restricted by a maximum pool size *c*.

| $c = 10$ | | | $c = 5$ | | |
| --- | --- | --- | --- | --- | --- |
| Pooling protocol | Prevalence Interval (%) |  | Pooling protocol | Prevalence Interval (%) |  |
|  | Lower bound | Upper Bound |  | Lower bound | Upper Bound |
| $S_{10}$ | 0.00 | 1.25 | $S_5$ | 0.00 | 6.625 |
| $S_9$ | 1.25 | 1.375 | $S_4$ | 6.625 | 12.375 |
| $S_{10 \times 10}$ | 1.375 | 4.875 | $S_3$ | 12.375 | 25.00 |
| $S_{9 \times 10}$ | 4.875 | 5.25 | | | |
| $S_{9 \times 9}$ | 5.25 | 5.875 | | | |
| $S_{8 \times 9}$ | 5.875 | 6.375 | | | |
| $S_{8 \times 8}$ | 6.375 | 7.375 | | | |
| $S_{7 \times 8}$ | 7.375 | 8.125 | | | |
| $S_{7 \times 7}$ | 8.125 | 9.625 | | | |
| $S_{6 \times 7}$ | 9.625 | 10.875 | | | |
| $S_{6 \times 6}$ | 10.875 | 12.125 | | | |
| $S_4$ | 12.125 | 12.375 | | | |
| $S_3$ | 12.375 | 25.00 | | | |

Prevalence intervals and their respective optimal pooling strategy. $S_m$ are single pooling protocols and $S_{m \times n}$ are matrix pooling protocols.

**Fig 6.**
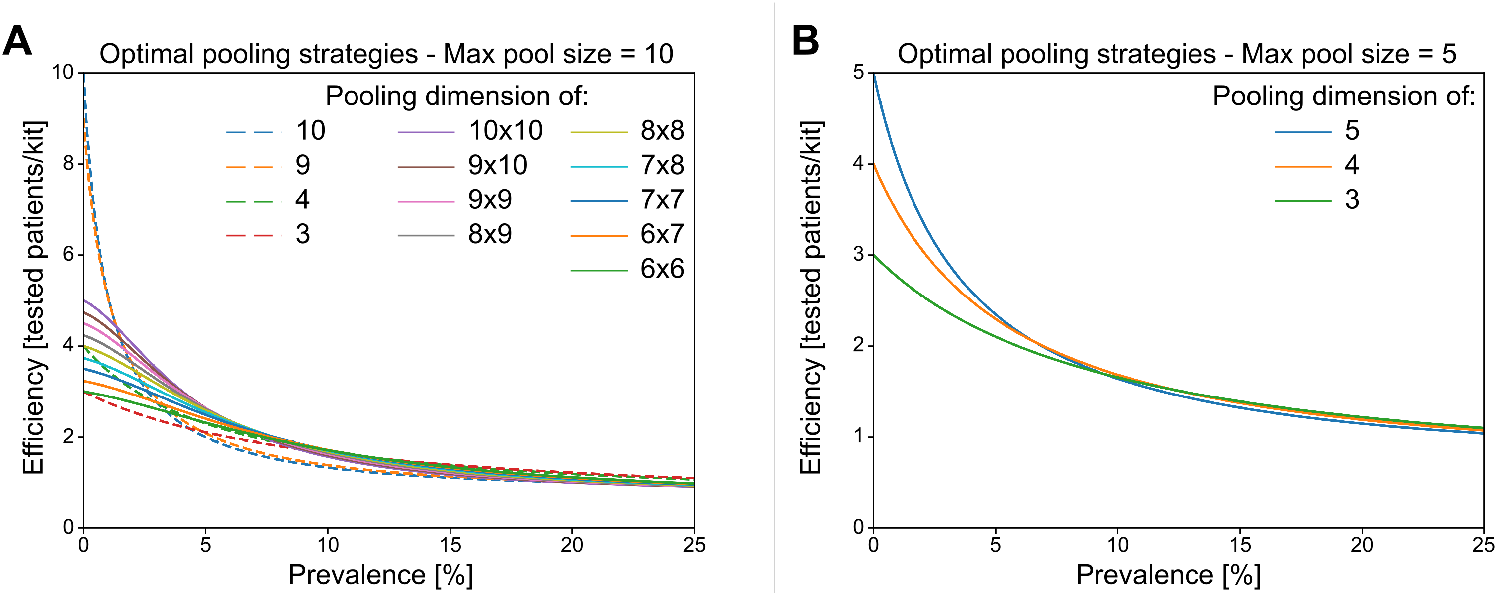
Optimal pooling strategies given a maximum pool size of **A**. 10 and **B**. 5.

## 5 Discussion

At the prevalence levels for COVID-19 that many diagnostic laboratories are currently managing, two-stage pooling efficiency is significantly reduced. Here, we show that it is possible to increase pooling efficiency by using machine learning to separate likely positive cases and then pool the rest of the samples using what we show to be an optimal strategy.

### Smart Pooling enhances but does not replace molecular testing

Smart Pooling uses artificial intelligence to enhance the performance of well-established diagnostics. It is an example of how data-driven models can complement, not replace, high-confidence molecular methods. Its robustness to the variability of the available data, prevalence, and model performance and its independence of pooling strategy, make it compelling to apply at large scales. Additionally, its continuous learning should make it robust to the pandemic’s evolution and our understanding of it.

### Smart Pooling could ease access to large scale testing

This pandemic has presented challenges to all nations, regardless of their income. As the number of infected people and the risk of contagion increase, more testing is required. However, the supply of test kits and reagents cannot cope with the demand, with most countries not being able to perform 0·3 new tests per thousand people [3]. Adopting Smart Pooling could translate into more accessible and larger-scale massive testing. In the case of Colombia, this could mean testing 50,000 samples daily, instead of 25,000, in mid-Jul 2020 [28]. If deployed globally, Smart Pooling truly has the potential to empower humanity to respond to the COVID-19 pandemic. It is an example of how artificial intelligence can be employed to bring social good.

## Data Availability

We will make our computational platform freely available for the general public in order to optimize COVID-19 testing.

## 6 Contributors

ME, GJ, LBS, AC, CG, DV, MFR, and JM designed and trained the AI models, and curated the dataset. MC, MGS, JMPL, and SR performed the molecular tests. ALM MFS, MV, and JMPL developed mathematical formulations to find the optimal pooling strategies. MFS and JMPL evaluated the applicability of the methods in the laboratory. ALM, MV and PA developed the theoretical interpretation of the results. LBS, GJ, JMW, JM, ALM, and MV prepared the figures. ME, GJ, LBS, AC, CG, DV, MFR, JM, JMW, MFS, and MV wrote the manuscript. All authors revised and accepted the final version of the manuscript. PA supervised and directed the research.

## 8 Data sharing

We will make our computational platform freely available for the general public in order to optimize COVID-19 testing, it is available indefinitely at the Smart Pooling website.

## 9 Acknowledgments

ME, GJ, and PA acknowledge special funding from Facultad de Ingeniería, Universidad de los Andes. The authors thank John Mario González from the Faculty of Medicine at Universidad de los Andes and his team CBMU for devoting the laboratory for COVID testing and their contributions during the laboratory certification. The authors also acknowledge the team members of the GenCore Covid Laboratory at Universidad de los Andes. The authors thank Secretaría de Salud de Bogotá and Alcaldía de Bogotá for their support during the laboratory certification process and access to complementary information for the samples.

## Supporting information

**Fig S1.**
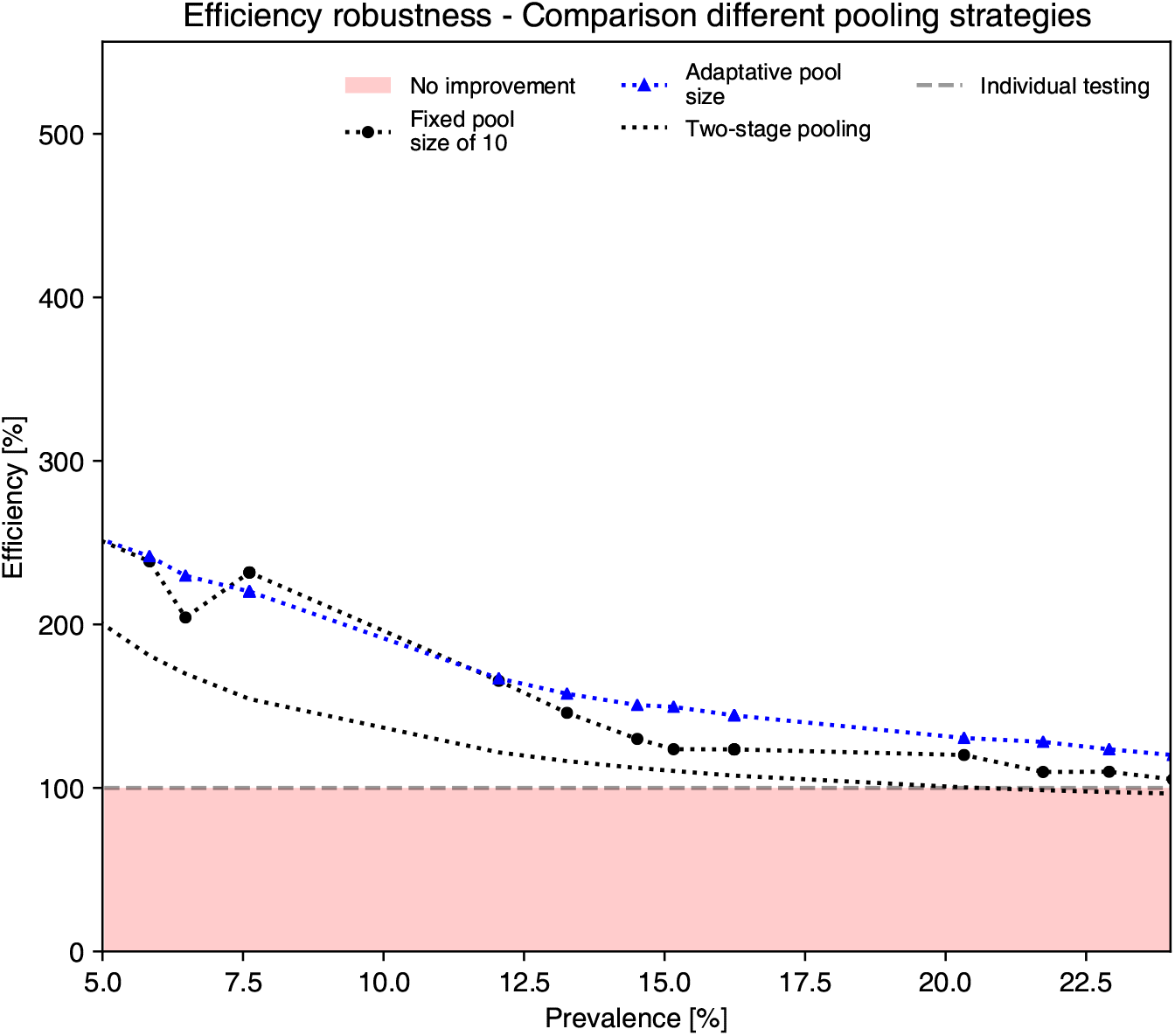
Estimated efficiency on the *Test Center Dataset* of different pooling strategies enhanced by Smart Pooling.

### S1.1 Practical considerations for implementation

Historically, pooling was used extensively during the second world war, but since then, it has mostly been implemented in specific niches such as testing blood for diseases and reducing cost in developing countries [18, 29, 30]. In the specific context of SARS-CoV-2, there are several incentives to implement pooling. Most significantly, the current shortage of reagents, especially in developing countries that do not produce these and have limited stocks. Additionally, if implemented correctly, these methods can increase throughput, another motivation for developing countries and locales with a limited number of certifiable qPCR machines. Finally, through the reduced use of reagents and increased throughput, these methods can reduce costs and motivate healthcare providers.

Here we considered Dorfman and array testing algorithms because they are easily compatible with a manual implementation of pooling. Although it is possible to find the optimal algorithm for pooling at a particular prevalence, it may be cumbersome to implement all the protocols. Fortunately, most of the pooling algorithms are relatively robust over a broader range than that in which they are optimal, so implementing some should increase efficiency without added complexity. Different algorithms will also affect tip use as well as plating time. Filter tips have been in short supply since the beginning of the pandemic [31], and some of the pooling protocols increase efficiency at the expense of increased tip use. Plating time will depend on both the pooling algorithm and the experience of the personnel doing the pooling. Each lab has to adapt the specifics of the protocol for their needs.

Successful implementation of PCR-based pooling requires understanding the limitations of the method and performing viability tests. Although it is possible to pool reactions that include primers that detect the virus only when it is present, it is not possible, to our knowledge, to pool the RNAse P positive control. This phenomenon occurs because the samples are positive unless a problem has occurred with the sample or the extraction of RNA; it involves finding a negative among positives where samples have variable RNA content and an exponential amplification step. In our implementation, the two reactions are separate. The RNAse P control is performed on a different plate, using a faster PCR protocol (1.5 hours instead of 2.5 hours). In kits for which the specific primers and the control are multiplexed, for example, it is not possible to ensure the presence of RNA when pooling.

Sensitivity must also be taken into account when implementing pooling. Each two-fold dilution results in the increase of the Ct value by 1 unit (1Δ*Ct*), on average. Some have detected dilutions up to 32-fold [9], but only for samples with average Ct, not for samples close to the detection limit. It is necessary to calibrate the dilution process in each lab since it depends on the kit, machine, and operators. Another limit for the size of the pools is the total reagent volume in each pool. Our plates can handle 10*µl* of a sample while other plate-kit combinations may handle just 5*µl*. Depending on the operators, pipetting volumes under 1*µl* may cause quality control problems. Since we rely on volunteers for our testing, we only considered pools of up to 10, which also keeps the maximum Ct of the first round below 42, the limit of cycles in our machine. Additionally, we increased the cutoff in Ct for the first round from 38 to 38+ Δ*Ct* at this dilution, from the calibrations. This modification may increase the number of false positives in the first round, but since the second round samples are sampled individually, the standard cutoff can be used to eliminate potential false positives from the first round.

Pooling before RNA extraction is an attractive option since it reduces reagent use, but we did not implement it for several reasons. The first is that the expected increase in Ct of 5 [32], was above the practical limits for our machine and kit combination. Secondly, it is not possible to flag poorly taken samples in which there is no RNA, for the same reasons, it is not possible to pool the RNAse P control. Finally, we received heterogeneous samples of the upper and lower respiratory tracts and when pooled together resulted in false negatives. It is likely that implementing pooling before RNA extraction is possible with other kits or more homogeneous samples.

The long-term management of COVID-19 will likely require the use of complementary approaches, including testing for antibodies against the virus. The methods presented in this paper could increase the measurement of seropositivity in the population.

## Footnotes

1 https://www.kaggle.com/rohanrao/covid-19-w3-lgb-mad/notebook

## References

1. Zhu N, Zhang D, Wang W, Li X, Yang B, Song J, et al. A novel coronavirus from patients with pneumonia in China, 2019. New England Journal of Medicine. 2020 feb;382(8):727–733.

2. World Health Organization. Coronavirus disease 2019 (COVID-19): situation report, 72. 2020;.

3. Max Roser EOO Hannah Ritchie, Hasell J. Coronavirus Pandemic (COVID-19). Our World in Data. 2020; https://ourworldindata.org/coronavirus.

4. Clark A, Jit M, Warren-Gash C, Guthrie B, Wang HHX, Mercer SW, et al. Global, regional, and national estimates of the population at increased risk of severe COVID-19 due to underlying health conditions in 2020: a modelling study. The Lancet Global Health. 2020; Available from: http://www.sciencedirect.com/science/article/pii/S2214109X20302643.

5. Dorfman R. The Detection of Defective Members of Large Populations. The Annals of Mathematical Statistics. 1943; 14(4):436–440.

6. Phatarfod RM, Sudbury A. The use of a square array scheme in blood testing. Statistics in Medicine. 1994; 13(22):2337–2343.

7. Kendziorski C, Irizarry RA, Chen KS, Haag JD, Gould MN. On the utility of pooling biological samples in microarray experiments. Proceedings of the National Academy of Sciences. 2005; 102(12):4252–4257. Available from: www.pnas.orgcgidoi10.1073pnas.0500607102.

8. Abdalhamid B, Bilder CR, McCutchen EL, Hinrichs SH, Koepsell SA, Iwen PC. Assessment of Specimen Pooling to Conserve SARS CoV-2 Testing Resources. American Journal of Clinical Pathology. 2020; 153:715–718.

9. Yelin I, Aharony N, Shaer-Tamar E, Argoetti A, Messer E, Berenbaum D, et al. Evaluation of COVID-19 RT-qPCR test in multi-sample pools. medRxiv. 2020; p. 2020.03.26.20039438. Available from: https://www.medrxiv.org/content/10.1101/2020.03.26.20039438v1.

10. Eis-Hübinger AM, Hönemann M, Wenzel JJ, Berger A, Widera M, Schmidt B, et al. Ad hoc laboratory-based surveillance of SARS-CoV-2 by real-time RT-PCR using minipools of RNA prepared from routine respiratory samples. Journal of Clinical Virology. 2020 jun;127.

11. Hogan CA, Sahoo MK, Pinsky BA. Sample Pooling as a Strategy to Detect Community Transmission of SARS-CoV-2. JAMA. 2020; 323(19):1967–1969.

12. Ghosh S, Rajwade A, Krishna S, Gopalkrishnan N, Schaus TE, Chakravarthy A, et al. Tapestry: A Single-Round Smart Pooling Technique for COVID-19 Testing. medRxiv. 2020;.

13. Randhawa GS, Soltysiak MP, El Roz H, de Souza CP, Hill KA, Kari L. Machine learning using intrinsic genomic signatures for rapid classification of novel pathogens: COVID-19 case study. PLoS ONE. 2020; 15(4):e0232391.

14. Gozes O, Frid-Adar M, Greenspan H, Browning PD, Zhang H, Ji W, et al. Rapid AI development cycle for the coronavirus (COVID-19) pandemic: Initial results for automated detection & patient monitoring using deep learning CT image analysis. 200305037. 2020;.

15. Yan L, Zhang HT, Goncalves J, Xiao Y, Wang M, Guo Y, et al. An interpretable mortality prediction model for COVID-19 patients. Nature Machine Intelligence. 2020; 2:283–288.

16. Petropoulos F, Makridakis S. Forecasting the novel coronavirus COVID-19. PLoS ONE. 2020; 15(3):e0231236.

17. Menni C, Valdes AM, Freidin MB, Sudre CH, Nguyen LH, Drew DA, et al. Real-time tracking of self-reported symptoms to predict potential COVID-19. Nature Medicine. 2020 may;Available from: http://www.ncbi.nlm.nih.gov/pubmed/32393804.

18. Taylor SM, Juliano JJ, Trottman PA, Griffin JB, Landis SH, Kitsa P, et al. High-throughput pooling and real-time PCR-based strategy for malaria detection. Journal of Clinical Microbiology. 2010; 48(2):512–519.

19. Weinberg CR. Editorial: Making the Best Use of Test Kits for COVID-19. American Journal of Epidemiology. 2020; Available from: https://academic.oup.com/aje/advance-article/doi/10.1093/aje/kwaa080/5831425.

20. Kucharski AJ, Klepac P, Conlan A, Kissler SM, Tang M, Fry H, et al. Effectiveness of isolation, testing, contact tracing and physical distancing on reducing transmission of SARS-CoV-2 in different settings. The Lancet. 2020; Available from: https://www.thelancet.com/journals/laninf/article/PIIS1473-3099(20)30457-6/fulltext.

21. Corman VM, Landt O, Kaiser M, Molenkamp R, Meijer A, Chu DKW, et al. Detection of 2019 novel coronavirus (2019-nCoV) by real-time RT-PCR. Eurosurveillance. 2020 jan;25(3).

22. SEASUN Biomaterials. U-TOP™ COVID-19 Detection Kit. USA Food and Drug Administration; 2020. Available from: https://www.fda.gov/media/137425/download.

23. H20ai. Python Interface for H2O; 2020. Python module version 3.10.0.8. Available from: https://github.com/h2oai/h2o-3.

24. Natekin A, Knoll A. Gradient Boosting Machines, a Tutorial. Frontiers in Neurorobotics. 2013; 7:21.

25. Hwang F. A Generalized Binomial Group Testing Problem. Journal of the American Statistical Association. 1975; 70(352):923–926.

26. Witt G. A simple distribution for the sum of correlated, exchangeable binary data. Communications in Statistics - Theory and Methods. 2014; 43(20):4265–4280.

27. Mutesa L, Ndishimye P, Butera Y, Uwineza A, Rutayisire R, Musoni E, et al. A strategy for finding people infected with SARS-CoV-2: optimizing pooled testing at low prevalence. arXiv preprint 200414934. 2020;.

28. de Salud IN. Coronavirus (COVID - 2019) en Colombia. 2020; https://www.ins.gov.co/Noticias/Paginas/Coronavirus.aspx.

29. Emmanuel J, Bassett M, Smith H, Jacobs J. Pooling of sera for human immunodeficiency virus (HIV) testing: an economical method for use in developing countries. Journal of Clinical Pathology. 1988; 41(5):582–585.

30. Quinn TC, Brookmeyer R, Kline R, Shepherd M, Paranjape R, Mehendale S, et al. Feasibility of pooling sera for HIV-1 viral RNA to diagnose acute primary HIV-1 infection and estimate HIV incidence. Aids. 2000; 14(17):2751–2757.

31. Cnn, Devine C, Griffin D, Kuznia R. Shortage of standard health supplies is ‘a huge problem’;. Library Catalog: edition.cnn.com. Available from: https://www.cnn.com/2020/03/18/us/coronovirus-testing-supply-shortages-invs/index.html.

32. Torres I, Albert E, Navarro D. Pooling of nasopharyngeal swab specimens for SARS-CoV-2 detection by RT-PCR. Journal of Medical Virology. 2020;.

